# The Association of Advanced Care Planning and Good Death in Palliative Elderly Acute Myeloid Leukemia Patients in Thailand

**DOI:** 10.1101/2024.03.18.24304500

**Authors:** Songphol Tungjitviboonkun

## Abstract

**Background:** The goal of treatment for elderly AML patients is palliative care rather than curative chemotherapy. Advanced care planning (ACP) plays a crucial role in good death. However, the factors associated with ACP and good death have not been well described.

**Objective:** This study aimed to characterize the association between advanced care planning and the outcome of good death in elderly AML patients.

**Method:** AML patients aged ≥60, who received less intensive treatment from August 2020 to December 2021, were interviewed and prospectively followed. All clinical data and potential factors related to ACP and good death were collected.

**Results:** Seventeen AML patients were included during the study period. The median age was 73 years (range 63-88). Twenty-five patients had passed away, with a median survival of 5.5 months. Thirteen patients had undergone ACP. Among them, twelve patients with ACP had passed away, and all had achieved good death, while six out of fifteen in the non-ACP group had achieved good death (p=0.003). Two-thirds of the ACP group initially expected to live for a year when undergoing ACP but later revised their expectations downward. In the non-ACP group, only two patients who achieved a good death were aware that AML would be the cause of death, whereas five out of six in the non-good death group were not aware of it. However, it was found that prognosis understanding alone did not correlate with successful end-of-life care. No other significant factors associated with patient outcomes were identified. Factors associated with the initiation of ACP discussions included decreased platelet count and increased blast count from baseline. Palliative performance status was not predictive when assessed too late. The median time from palliative consultation to death was 3.5 months, which appeared sufficient for patient preparation.

**Conclusion:** The key factor associated with good death was the initiation of ACP discussions with the patient. Decreased platelet count and increased blast count from baseline could serve as warning signs.

## Introduction

Up to 70 percent of Acute Myeloid Leukemia (AML) cases occur in people over 60 years of age. Although new treatments are available, the death rate is still high and the response rate to chemotherapy is low [1]. The decision to treat AML patients older than 60 years requires careful and comprehensive patient evaluation, including the health of the patient, the disease, psychological and social readiness, and economic status as well as having someone to take care of and planning the treatment together with the patient and relatives. Treatment approaches can be 1) intensive treatment, the goal is a complete cure; 2) less intensive treatment, the goal is to prolong the survival 3) Palliative treatment, treatment according to the symptoms that the patient has from the disease [2].

Palliative care is holistic health care including physical, mental, social, and spiritual aspects, so that patients can face illness through prevention processes and relieve suffering from pain. The main goal is to increase the best quality of life in the remaining time according to the context of the individual.

Advance care planning is an important part of providing palliative care. To provide care for terminally ill patients so that they pass away peacefully with minimal suffering, maintaining human dignity, and reducing conflict between relatives and the health team about how each type of life-sustaining procedure or treatment is appropriate. This has resulted in an effort from all parties involved to develop a care model for advanced planning in the same direction.[3, 4]

The term “good death” has been studied by many people. The first and most commonly cited report came from the Health and Medicine Division, the National Academies of Sciences, Engineering, and Medicine, USA (formerly the Institute of Medicine), in 1997, providing a definition of a good death is “one that is free from avoidable distress and suffering for patient, family, and caregivers, in general accord with the patient’s and family’s wishes, and reasonably consistent with clinical, cultural, and ethical standards.” A death that is free from worry or avoidable suffering of the patient’s family and caregivers should generally be based on the needs of the patient and family, reasonably consistent with the symptom’s cultural standards and ethics [5, 6]

In 2016, Meier and colleagues reported on a review of research on the definition of a good death or successful dying across patient groups, families, and medical personnel, a total of 36 studies, found definitions to be divided into the top 3 topics as follows [7]

1. Preferences for the dying process: means the deceased accepts and is ready to leave. The death scene: how, who, where, and when is planned.
2. Pain-free status: means death without pain or suffering
3. Emotional well-being: means receiving emotional and spiritual care as needed.

Dying well is an important goal. To assess the quality of end-of-life care, there are various tools used to assess good death, but there is still no standard and no form for evaluating good death in Thailand. Good death assessment forms that are frequently referenced include: The Quality of Dying and Death (QODD) questionnaire [8], and Good Death Inventory (GDI) [9]. When considering the details of these assessment, it cannot be used in the study because some questions do not fit the social situation in Thailand. There were difficulties in using and interpreting the results, so the questions were modified, regrouped, and consulted with end-of-life care expert team. To cover the desired aspect of a good death and appropriate to the situation of patients in the hospital. Therefore, a new model for assessing a good death was created specifically for use in this study.

## Materials and Methods

### Objectives

Primary objective is to know the results of advanced care plan to the outcome of good death in elderly patients with AML assessed from end-of-life records. Secondary objectives are to assess the quality of care by measuring both patient quality of life during follow-up and stress of relatives during follow-up.

### Method

This study conduct prospective observational descriptive study research in patients with AML aged ≥ 60 years who receive palliative treatment at KCMH. Either new diagnosis or continued follow-up from 1^st^ August 2020 to 31^st^ December 2021.

### Primary outcome

Good death of elderly patients with AML who receive palliative care and have advanced care planning compare to who did not have advanced care planning.

### Secondary outcome

The study collected data on patients’ quality of life, relatives’ stress, advanced care planning, end-of-life care, and when doctors initiate discussions about advanced care planning. The researcher met with patients and relatives separately, providing different explanations and documents to ensure sensitive messages were handled appropriately. Patients and relatives had the opportunity to ask questions and make independent decisions upon signing a voluntary consent form. In cases where the researcher was directly involved in patient care, a co-researcher handled the consent process.

Patients received standard medical care from their doctors, while the research team gathered information on care and the emotions of patients and relatives. Personal interviews were conducted separately with patients and relatives after each patient’s appointment with the attending doctor. These interviews occurred at the start of the study and every 8 weeks thereafter, continuing until the patient’s passing.

In the event of a patient’s death, end-of-life characteristics were recorded within 48 hours, and a ‘good death’ evaluation was conducted. The evaluation involved three medical personnel: the treating doctor, a caring nurse, and a palliative care specialist. To maintain objectivity, evaluators were unaware of the patient’s advanced care planning status. If the patient passed away elsewhere, we conducted telephone interviews with relatives using the same questions.

A ‘good death’ assessment considered the patient’s preferences for the dying process, freedom from pain, and emotional well-being. A patient was categorized as having had a ‘good death’ when at least 2 out of 3 evaluators agreed on this classification.

## Research tools

Consists of 5 parts as follows:

### Part 1

Palliative Performance Scale (PPS version 2) assessment form, translated by the palliative care committee, Nursing Department, Maharaj Nakhon Chiang Mai Hospital, on May 13, 2008. The Palliative Performance scale was developed in 1996 by Anderson and colleagues at the Victoria Hospice Society (VHS) as a tool for assessing patient conditions.

### Part 2

Quality of life assessment using the FACT-Leu form for patients with Leukemia[10].

### Part 3

Relative stress assessment form adapted from a distress thermometer.

### Part 4

Advanced care planning for patients and relatives, created by the researcher.

### Part 5

Form for recording the last moments of life, used to assess whether it constitutes a ‘good death.’ This form is based on Meier et al.’s definition and adapted from the Good Death Inventory (GDI), focusing on clear indicators for patients’ preferences for the dying process, pain-free status, and emotional well-being.

## Data Collection

Data collection includes gathering general patient information such as age, gender, occupation, religion, and medical history, along with details about their living situation, caregivers, and a palliative performance scale evaluation. Additionally, data is collected on close relatives, clinical aspects of the disease, treatment methods, hospital admissions, complete blood count results, and blood transfusion history. Palliative care information is recorded, as well as quality of life assessments, evaluations of relatives’ stress levels during and after the patient’s passing, and monitoring of caregiver stress. Advance care planning details are documented for both patients and relatives. End-of-life characteristics, including cause and place of death, individuals present during the final days, treatments received, resuscitation efforts, pain and discomfort symptoms at the time of death, involvement in spiritual or religious activities, family meetings, and support offered to cope with loss are recorded. Miscellaneous information includes hospital length of stay, expenses incurred, and an evaluation form to determine whether it was a ‘good death’ or not.

## Ethical consideration

The researcher is a trained doctor with expertise in delivering difficult news. They possess the necessary skills to communicate effectively, ensuring that the information shared does not have a detrimental impact on the emotional well-being of patients and their relatives.

This study was conducted in accordance with the ethical standards outlined by the Declaration of Helsinki and its subsequent amendments. All procedures performed involving human participants were approved by the Institutional Review Board of the Faculty of Medicine Chulalongkorn University prior. IRB No. 670/63. Consent was informed to all patients and guardians in both verbal and written type and informed consent was signed from all individual participants included in the study.

## Results

### Baseline characteristics

A total of 28 patients were included, no patients were excluded. Among elderly patients with AML receiving palliative care, 13 out of 28 patients (46%) underwent advanced care planning (ACP). The median age for patients in the ACP group was 73 years (range, 63 to 88 years), while those without ACP had a median age of 69 years (range, 61 to 84 years). There was no statistically significant difference in age between the two groups (p=0.70).

Regarding treatment plans, patients in the ACP group received hypomethylating agents in 8 cases, cytoreductive agents in 3 cases, induction chemotherapy in 1 case, and supportive care with blood transfusions or blood components in 1 case. In contrast, the non-ACP group received hypomethylating agents in 9 cases, cytoreductive agents in 5 cases, and palliative care through blood transfusions or blood components in 1 case. There were no significant differences in treatment plans between the two study groups (p=0.70) (Table 1).

**Table 1:**
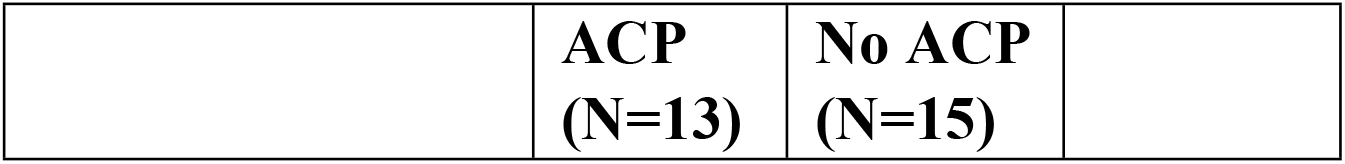
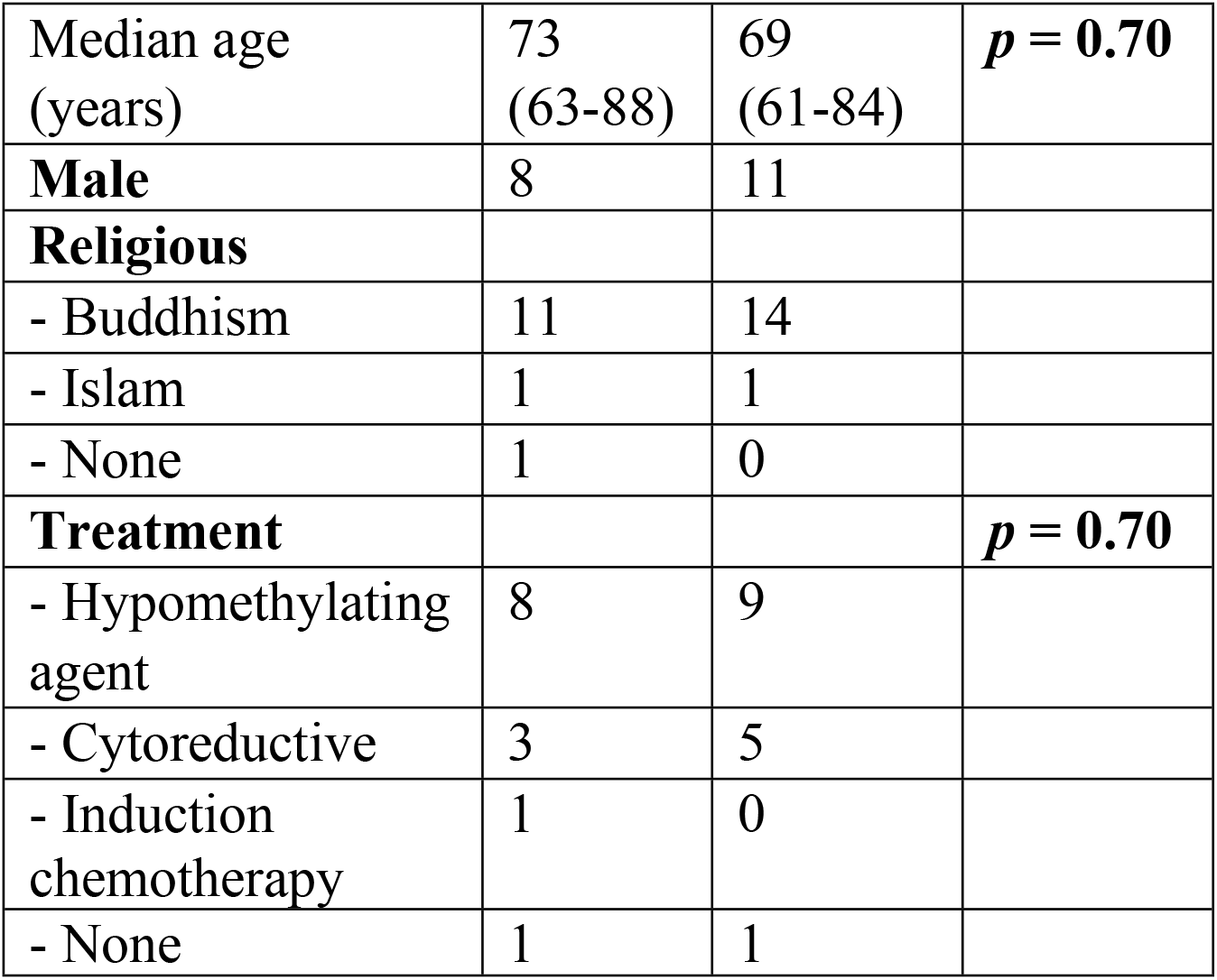
Baseline characteristics.

|  | ACP<br>(N=13) | No ACP<br>(N=15) |
| --- | --- | --- |

|  |  |  |  |
| --- | --- | --- | --- |
| Median age (years) | 73 (63-88) | 69 (61-84) | ***p* = 0.70** |
| **Male** | 8 | 11 |  |
| **Religious** |  |  |  |
| - Buddhism | 11 | 14 |  |
| - Islam | 1 | 1 |  |
| - None | 1 | 0 |  |
| **Treatment** |  |  | ***p* = 0.70** |
| - Hypomethylating agent | 8 | 9 |  |
| - Cytoreductive | 3 | 5 |  |
| - Induction chemotherapy | 1 | 0 |  |
| - None | 1 | 1 |  |

### Primary outcome

The primary outcome of the study revealed that among the 13 patients who underwent advanced care planning, 12 passed away during the study, and all 12 of them met the criteria for a “good death.” In contrast, among the 15 patients who did not engage in advanced care planning, 6 of the 13 who passed away during the study met the criteria for a “good death” (p=0.003) (Table 2).

**Table 2:** End-of-life characteristics.

|  | **ACP**<br>**(N=13)** | **No ACP**<br>**(N=15)** |  |
| --- | --- | --- | --- |
| **Death** | 12<br>(92.3%) | 13<br>(86.7%) |  |
| Good death | 12 | 6 | ***p* = 0.003** |
| Place of Death |  |  |  |
| - Home/ Local hospital | 7 | 2 | ***p* = 0.025** |
| - Tertiary hospital | 5 | 11 |  |
| Median time Diagnosis to Death (month) | 4.99<br>(1.77-16.77) | 7.92<br>(1.37-17.0) | ***p* = 0.695** |
| Median time Palliative to Death (month) | 1.5<br>(0-10.10) | 3.24<br>(0-7.83) |  |

Furthermore, patients who had advanced care planning tended to pass away either at home or in a nearby hospital (7 cases), while 5 patients in this group died at KCMH. Conversely, among those who did not have advanced care planning, a smaller number (2 cases) passed away at home or a nearby hospital compared to those with advanced care planning (p=0.025) (Table 2).

### Secondary outcome

In terms of secondary outcomes, patients who underwent advanced care planning had a median time from diagnosis to death of 4.99 months (range 1.77 to 16.80 months). On the other hand, patients who did not engage in advanced care planning had a median time from diagnosis to death of 7.92 months (range 0.50 to 17.07 months). The statistical analysis did not reveal a significant difference between the two study groups (p=0.695) (Figure 2).

**Figure 1:**
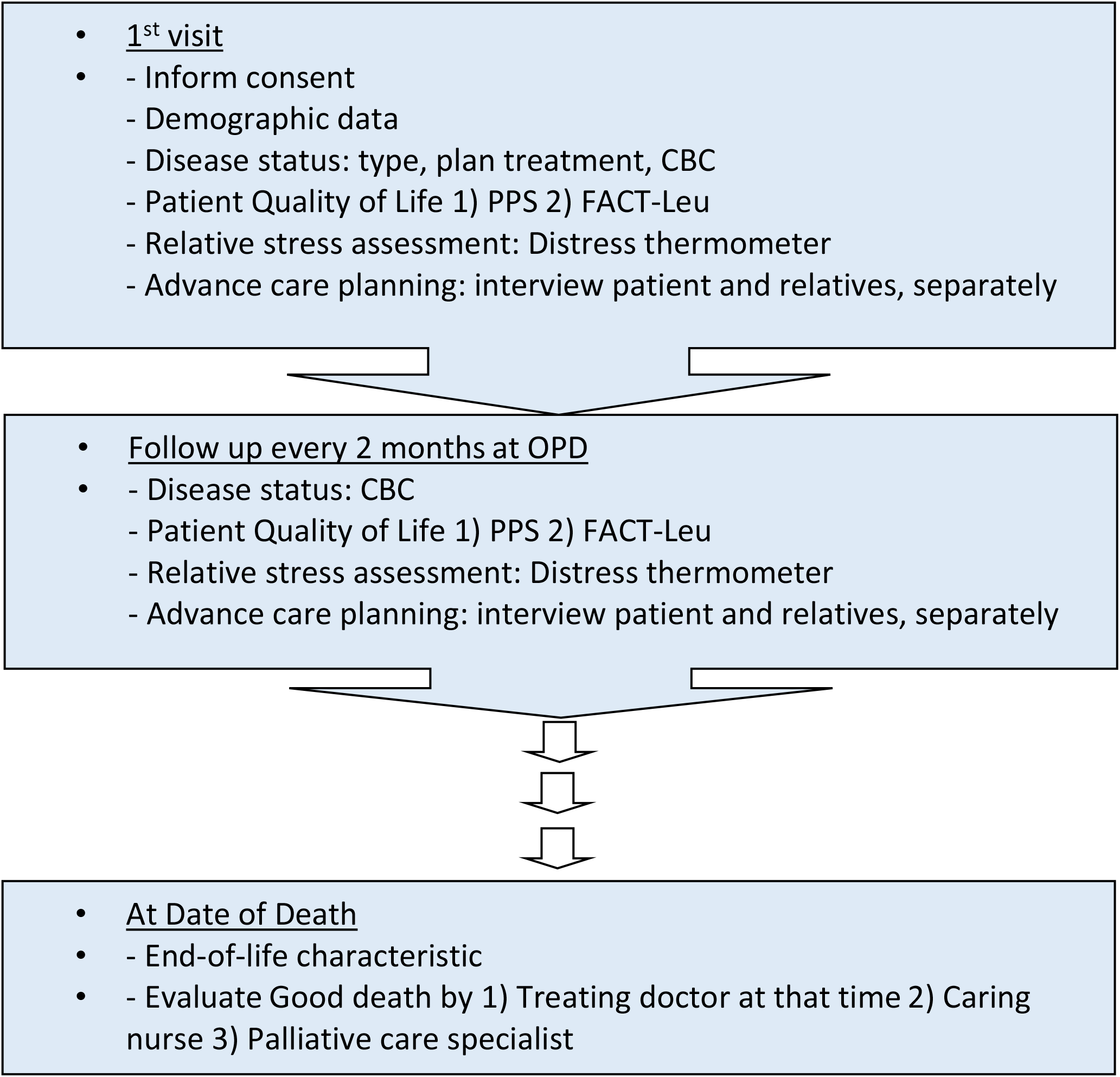
Flow chart for conducting research

**Figure 2:**
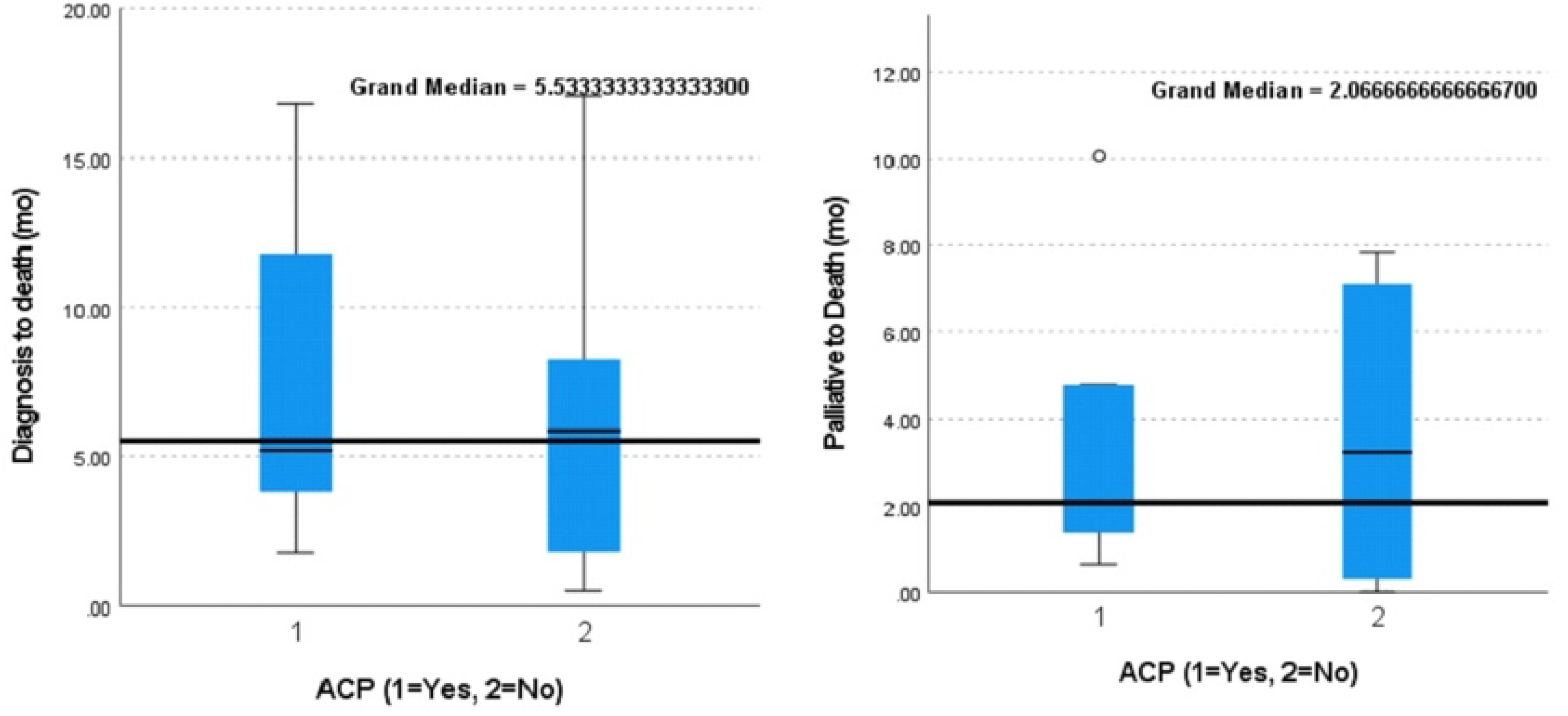
Compare the time from diagnosis to death. and the time from consultation with the palliative nurse or biomedical team until death. Comparison between 2 groups (Group 1 ACP group, Group 2 no ACP group)

In the groups that underwent advanced care planning, the stress levels of relatives, as measured by the Distress Thermometer, were lower compared to the group that did not have a care plan. Specifically, the group with advanced care planning had a median stress score of 2 points, while the group without advanced care planning had a higher median score of 5 points out of 10 (p=0.28) (Figure 3).

**Figure 3:**
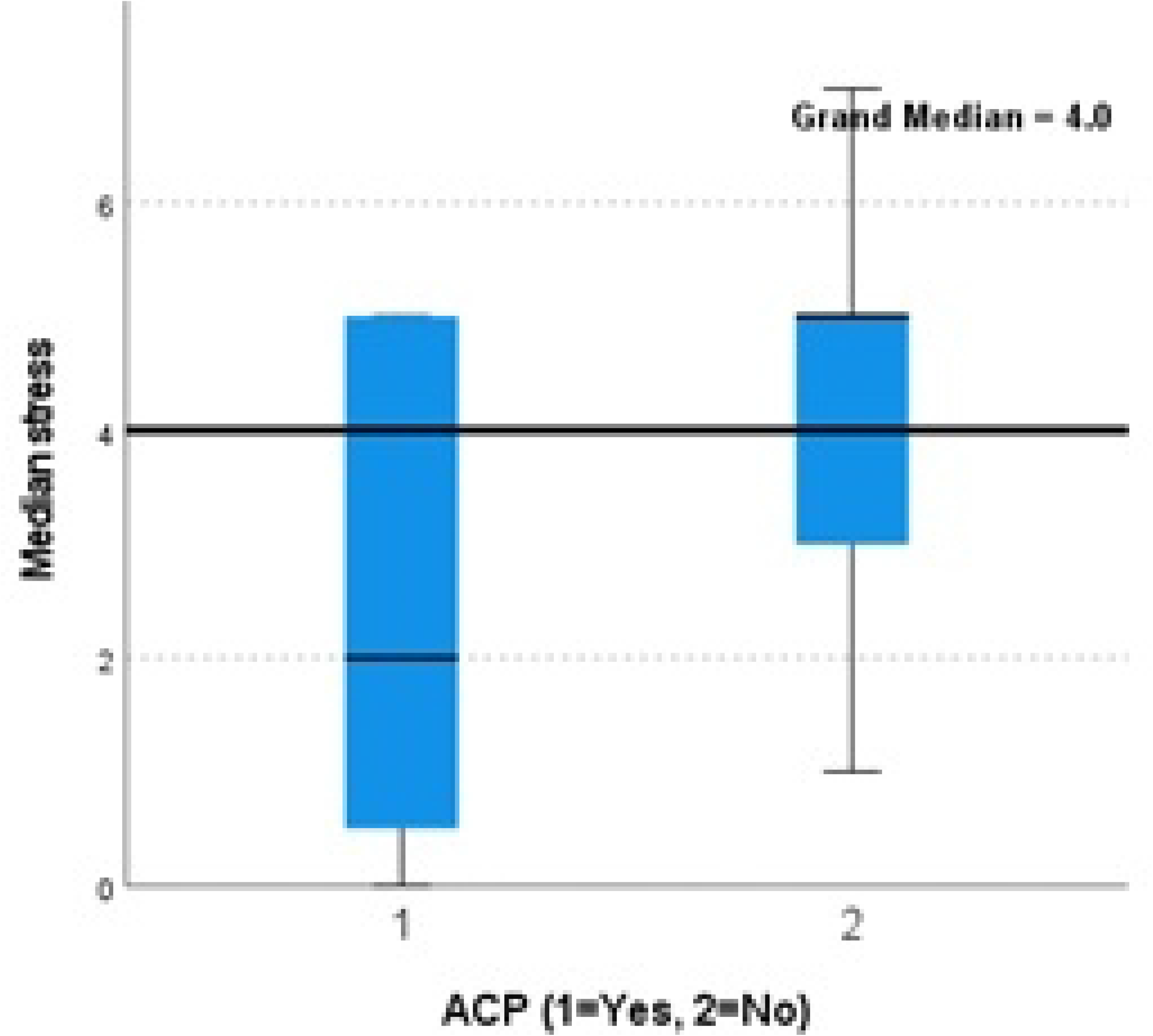
Compare relative stress scores between the 2 groups (Group 1 ACP group, Group 2 no ACP group).

The total hospital care costs from the time of diagnosis in the groups with advanced care planning appeared to be lower compared to those without plans. Specifically, the medians were 12,782 baht and 105,336 baht, respectively. However, this difference did not reach statistical significance (p=0.256) (Figure 4).

**Figure 4:**
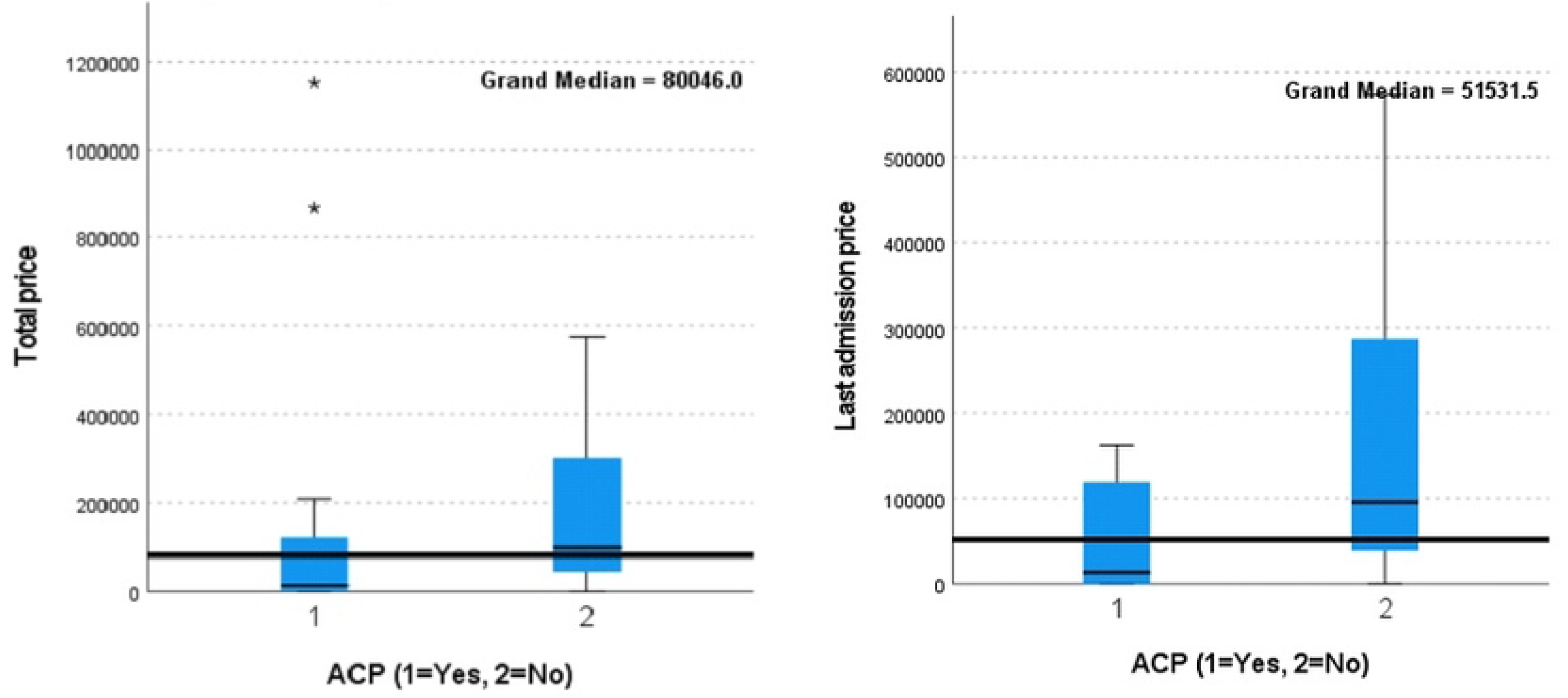
Compares total hospital care costs since diagnosis and hospital care costs only for the last hospital stay before death between the 2 groups (Group 1 ACP group, Group 2 no ACP group)

In contrast, when considering the cost of hospital care for the last hospital stay before death in the group that had advanced care planning, it was significantly lower than the group that did not have advanced care planning. The medians were 12,782 THB and 95,414 THB, respectively (p=0.058).

Regarding the factors influencing a doctor’s decision to initiate discussions about advanced care planning, it was observed that 7 out of 13 cases commenced these discussions when there was a significant deterioration in the patient’s blood results. This deterioration included a more than twofold increase in white blood cell count in 5 cases and a 50 percent decrease in platelets in 2 cases. These patients, on average, had a survival time of only 2 months following the change in blood results.

Among the 17 patients in the study, the attending physician initiated discussions about palliative care goals as early as 1 month from the time of diagnosis in 4 patients. Of these, three patients engaged in advanced care planning and experienced a good death, while one patient initiated discussions about treatment goals but did not have advanced care planning. Unfortunately, this patient passed away in the second month without achieving a good death.

## Discussion

This study highlights a significant association between advanced care planning and achieving a good death. Patients who engaged in advanced care planning exhibited better social and family well-being, emotional and mental states, and performed activities more satisfactorily compared to those without a care plan. Additionally, relatives of patients with advanced care plans experienced lower levels of stress.

Remarkably, advanced care planning did not lead to a shorter remaining life expectancy when compared to patients without such plans. The treatment plans developed by doctors remained consistent between the two groups. The primary difference lay in end-of-life care, debunking the previous misconception in some families that patients who are aware of their diagnosis may worsen their symptoms and die sooner.

The key distinction in end-of-life treatment was that patients with advanced care plans were more likely to pass away in their preferred setting, such as at home or in a nearby hospital, rather than a tertiary hospital. This suggests potential cost savings in care.

An essential factor motivating doctors to initiate discussions about advanced care planning is the observation of deteriorating leukemia conditions, often indicated by increased white blood cell counts and significantly reduced platelet levels. Patients in this category typically have an average of only 2 months left to live.

Therefore, advanced care planning is associated with achieving a good death without impacting other treatment outcomes and may even reduce costs. Doctors are encouraged to initiate advanced care planning discussions as early as possible, given the finding that patients often pass away sooner than expected. At the latest, these discussions should begin when there is a significant change in the patient’s white blood cell and platelet counts, indicating an estimated survival of approximately 2 months.

## Data Availability

All relevant data are within the manuscript and its Supporting Information files.

## Acknowledgement

The authors would like to express their gratitude to the Thai Society of Hematology for their generous funding support, which enabled the completion of this study.

